# Modified Rankin Scale Disability Status at Day 4 Poststroke is an Informative Predictor of Long-Term Day 90 Outcome

**DOI:** 10.1101/2023.06.30.23292102

**Authors:** Shayandokht Taleb, Jenny Ji-hyun Lee, Samuel Asanad, Sidney Starkman, Scott Hamilton, Robin A. Conwit, Nerses Sanossian, Jeffrey L Saver

**Affiliations:** Department of Neurology, David Geffen School of Medicine at UCLA; Department of Neurology, Kaiser Permanente Los Angeles Medical Center; Department of Ophthalmology and Visual Sciences, University of Maryland School of Medicine; Departments of Emergency Medicine and Neurology, David Geffen School of Medicine at UCLA; Department of Neurology, Stanford University; Department of Neurology, University of Indiana and NINDS Division of Clinical Research; Department of Neurology, Keck School of Medicine at USC

**Keywords:** Acute Cerebrovascular Disease, Cerebral Ischemia, Intracranial Hemorrhage, Disability, Outcome

## Abstract

**Background:** Long-term disability after stroke is standardly assessed 3 months post-onset, using the modified Rankin Scale (mRS). The value of an early, day 4 mRS assessment for projecting the 3-month disability outcome has not been formally investigated.

**Methods:** In this cohort of patients with acute cerebral ischemia and intracranial hemorrhage, we analyzed day 4 and day 90 mRS assessments in the NIH Field Administration of Stroke Therapy– Magnesium (FAST-MAG) Phase 3 trial. The performance of day 4 mRS, alone and as part of multivariate models, in predicting day 90 mRS was assessed using correlation coefficients, percent agreement, and the kappa statistics.

**Results:** Among the 1573 acute cerebrovascular disease (ACVD) patients, 1206 (76.7%) had acute cerebral ischemia (ACI), while 367 (23.3%) had intracranial hemorrhage. Among all 1573 ACVD patients, day 4 mRS and day 90 mRS correlated strongly, Spearman’s rho=0.79, in unadjusted analysis with weighted kappa of 0.59. For dichotomized outcomes, simple carry-forward of the day 4 mRS performed fairly well in agreeing with day 90 mRS: mRS 0-1 (k=0.67), 85.4%; mRS 0-2 (k=0.59), 79.5%; fatal outcome, 88.3% (k=0.33). Correlations of 4d and 90d mRS were stronger for ACI than ICH patients, 0.76 vs 0.71.

**Conclusions:** In this acute cerebrovascular disease patient cohort, assessment of global disability performed on day 4 is highly informative regarding long-term, 3-month mRS disability outcome, alone, and even more strongly in combination with baseline prognostic variables. The day 4 mRS is a useful measure for imputing the final patient disability outcome in clinical trials and quality improvement programs.

## Introduction

Stroke is a disease that disables more often that it kills, and assessment of post-stroke disability is crucial to the evaluation of the efficacy of novel treatments in clinical trials and the effectiveness of conventional care in standard practice.^1, 2^ Three months is generally recognized as the most appropriate time point for assessing functional outcomes after an acute stroke.^3, 4^ By 3 months, most of the recovery in function due to neuroplasticity and neurorepair has transpired. While additional recovery may occur in some patients over the rest of the first post-stroke year, and beyond, as more time progresses, recurrent strokes and other competing causes of morbidity and mortality are more likely to arise, interfering with direct assessment of the effects of the index stroke. The 3-month time frame balances these contending forces to provide the most informative assessment of post-stroke functional state. Consequently, it is the time frame most frequently chosen for measuring primary outcomes in acute stroke trials and quality improvement programs.

However, a challenge to the use of a 3-month assessment is that patients may be lost to follow-up in the long interval after their discharge from the acute hospitalization, especially in a mobile society with a fragmented system of medical care.^5–7^ To analyze clinical trial results and to guide quality improvement programs, it is important to develop valid methods of imputing 3-month outcomes for patients who do not have direct assessments available. The most widely used outcome measure is the modified Rankin Scale (mRS), a seven-level global assessment of combined disability and handicap.^4^ Prior studies have found that earlier mRS values, obtained at day 7 or at day 30, are the single most important predictor of the mRS value at day 90, and have provided multivariate formula for predicting day 90 mRS that incorporate these earlier mRS values.^8, 9^ However, systems of care have changed since the studies from which those equations were derived, and the median length of stay for an acute stroke hospitalization is now substantially less than in the past.^10^ For acute ischemic stroke, the median length of stay is 4-5 days, and even day 7 mRS assessments would take place after discharge for most patients, when the risk for loss to follow-up is higher.^10, 11^ In contrast, day 4 mRS values are more reliably available, as many patients are still in the acute hospital on this date and the remaining patients are only recently discharged and generally still motivated to be accessible. Predictive models for day 90 mRS that incorporate the day 4 mRS would therefore further address data missingness in clinical trials and quality improvement programs. Such a very early assessment would also be of value as a dependable early endpoint to drive randomization adjustments in clinical trials using adaptive dose-adjustment designs, ^12, 13^ and clinically helpful in providing patients and families with long-term prognoses and home care planning.

## Methods

We analyzed all patients with acute cerebral ischemia and intracranial hemorrhage with both day 4 and day 90 mRS assessments in the database of the NIH Field Administration of Stroke Therapy– Magnesium (FAST-MAG) Phase 3 trial. At both time points, the mRS was assessed by certified nurse study-coordinators, in person, or telephonically if patient and caregivers were accessible only by phone.

Since the primary trial results showed a neutral effect of magnesium sulfate on outcome, without an interaction effect between treatment group and mRS score, we combined the placebo and magnesium groups in the analysis. Missing data were handled using complete case analysis.

For the day 90 mRS, raters were required to score the mRS using a formal structured assessment tool – the Rankin Focused Assessment (RFA).^14^ For the day 4 mRS, raters were encouraged but not required to use the RFA. In addition, rater instructions for the day 4 mRS were that the score be based on the rater’s judgment of what the subject was capable of doing on day 4, using not only patient report, but also available information from family, physicians and nurses, physical and occupational therapists, and the rater’s own direct assessment including an NIH Stroke Scale and a Barthel Index evaluation. Raters were directed that the scores should indicate their best judgment of what activities the patient could do on day 4, not simply what the patients had had an opportunity to do so far. For example, if a patient had fully recovered but remained in hospital solely for diagnostic tests, the mRS was scored indicating the patient was work-capable.

The performance of the day 4 mRS full ordinal 7 levels in predicting day 90 full ordinal 7 mRS levels was assessed using correlation coefficients, percent agreement, and the weighted kappa statistic, including both unadjusted and multivariate adjusted analyses. The performance of the day 4 mRS levels in predicting day 90 mRS dichotomized outcomes was assessed for freedom-from-disability (mRS 0-1) and functional independence (mRS 0-2), using correlation coefficients, percent agreement, and kappa statistics, including both unadjusted and multivariate adjusted analyses. Kappa values were considered as indicating: 0.00-0.20 none to slight; 0.21-0.40 fair, 0.41– 0.60 moderate, 0.61- 0.80 substantial, and 0.81-1.00 almost perfect agreement.

The multivariate predictive models examined the predictive ability of the day 4 mRS and other clinically relevant predictor variables including NIHSS and age for all the models. The models examined the predictive ability of 17 additional demographic and clinical covariates, which were selected based on prior studies. The variables with the minimum Akaike Information Criterion (AIC) were purposefully included in the model. These candidate predictor covariates included 4 additional continuous variables: body mass index, pre-stroke mRS, admission serum glucose, and initial CT/MRI ASPECTS score. The model also included 13 dichotomized or categorical variables: sex, race, Hispanic ethnicity, stroke subtype (acute cerebral ischemia versus intracranial hemorrhage), hypertension, diabetes, dyslipidemia, atrial fibrillation, smoking (current or within past year), myocardial infarction or coronary artery disease, prior symptomatic cerebrovascular disease, any current alcohol use, and concomitant alteplase treatment.

## Results

Among the 1700 patients enrolled in the FAST-MAG trial, 1244 patients had transient ischemic attack or acute ischemic stroke, while 387 patients had intracranial hemorrhage (ICH). A total of 67 were excluded from the current analysis study due to having a stroke-mimicking condition as the final diagnosis for their presenting event, 42 were excluded for not having documented modified Rankin Scale (mRS) scores at both day 4 and day 90. Accordingly, univariate analyses were conducted on 1573 patients, acute cerebrovascular disease (ACVD) patients with documented mRS scores at both day 4 and day 90, including 1206 (76.7%) with acute cerebral ischemia (ACI) and 367 (23.3%) with acute ICH. The characteristics of these patients are shown in Table 1. The unadjusted distributions of day 4 and day 90 mRS scores for patients with ACI and ICH are shown in Table 2 with more severe disability distributions for the patients with ICH at both timepoints.

**Table 1.**
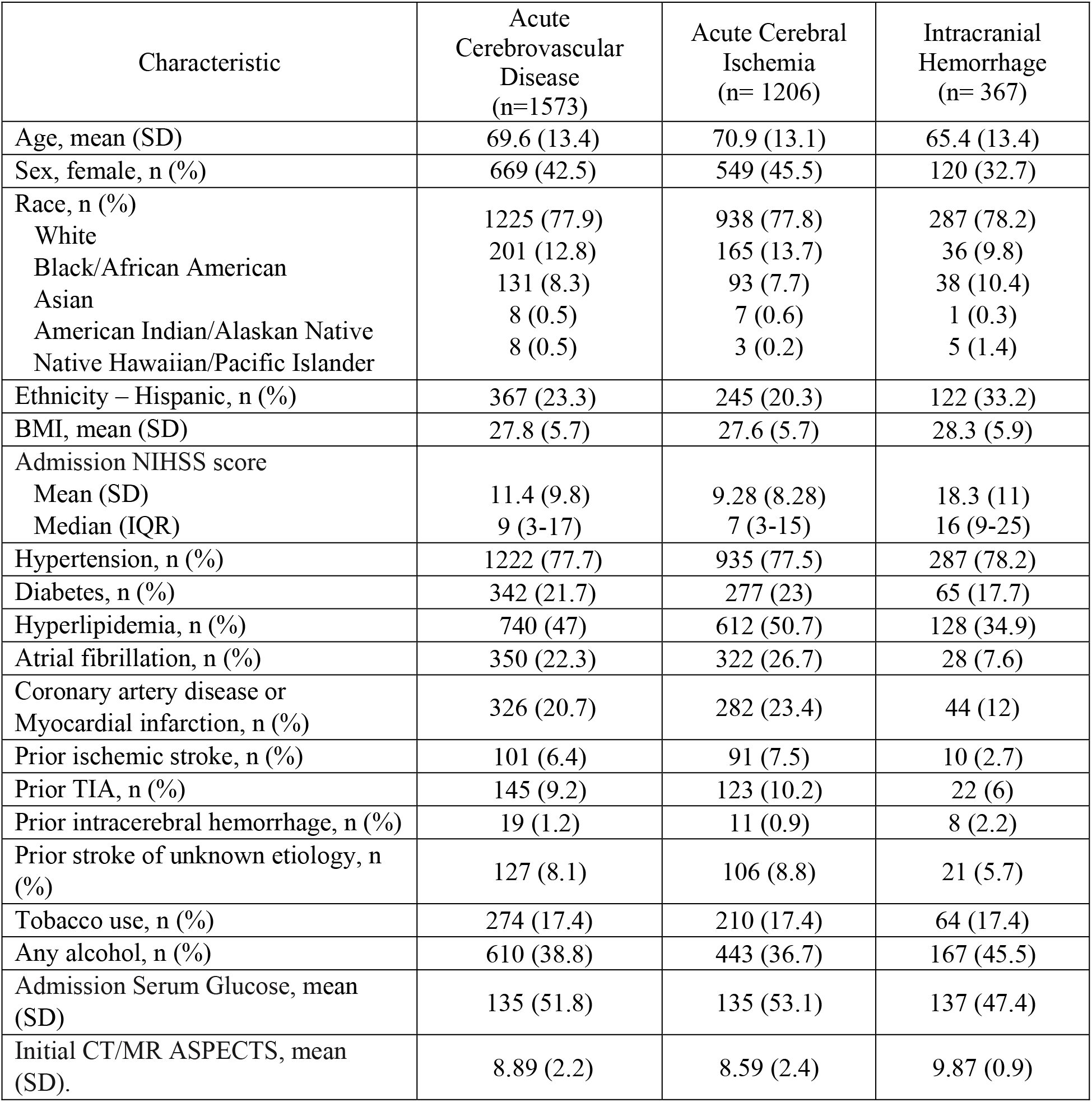
Baseline Characteristics of Study Patients

**Table 2.**
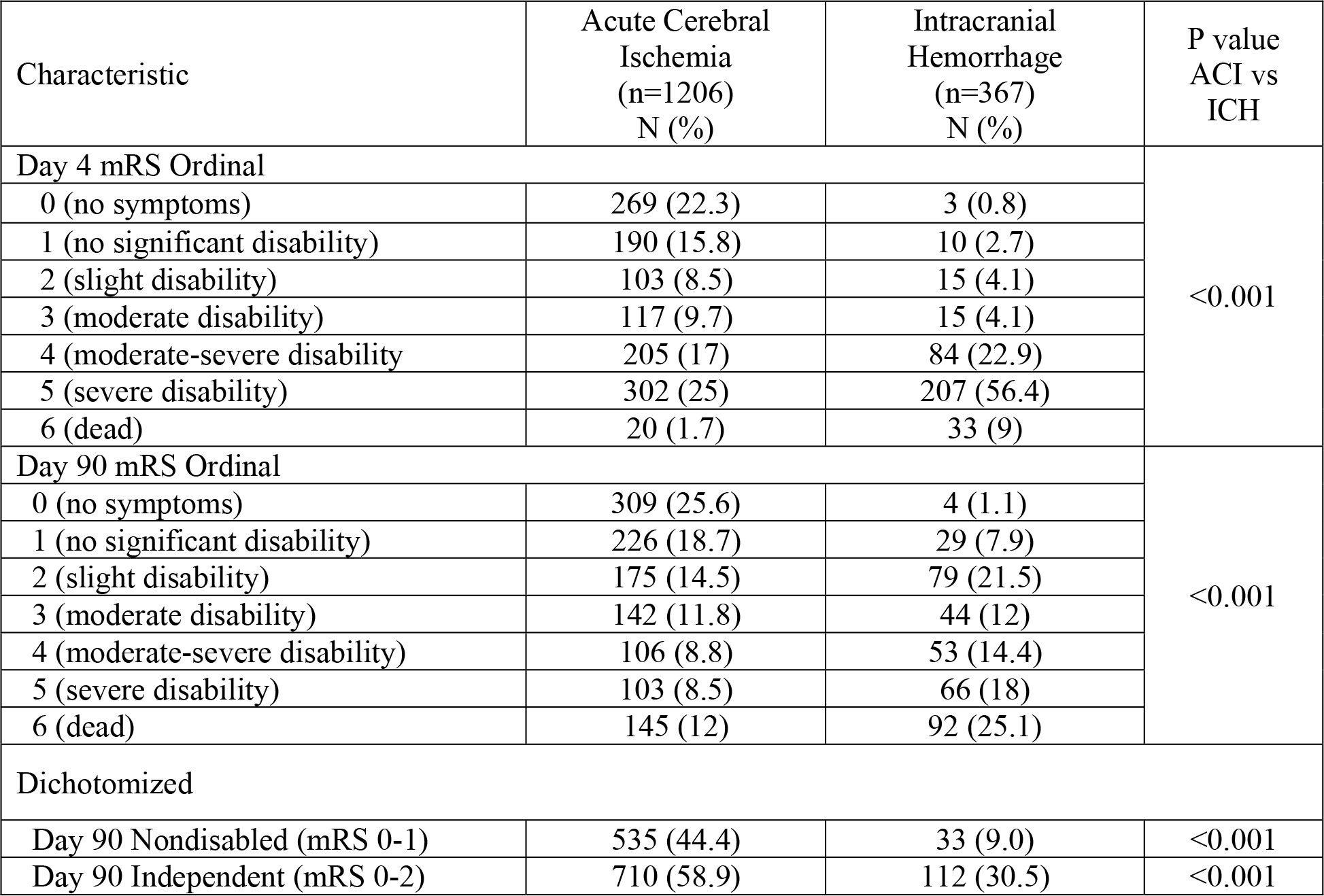
Day 4 mRS and Day 90 Distributions Among Acute Cerebral Ischemia and Intracranial Hemorrhage Patients

| Characteristic | Acute Cerebral Ischemia<br>(n=1206)<br>N (%) | Intracranial Hemorrhage<br>(n=367)<br>N (%) | P value<br>ACI vs ICH |
| --- | --- | --- | --- |
| Day 4 mRS Ordinal |  |  | <0.001 |
| 0 (no symptoms) | 269 (22.3) | 3 (0.8) |  |
| 1 (no significant disability) | 190 (15.8) | 10 (2.7) |  |
| 2 (slight disability) | 103 (8.5) | 15 (4.1) |  |
| 3 (moderate disability) | 117 (9.7) | 15 (4.1) |  |
| 4 (moderate-severe disability) | 205 (17) | 84 (22.9) |  |
| 5 (severe disability) | 302 (25) | 207 (56.4) |  |
| 6 (dead) | 20 (1.7) | 33 (9) |  |
| Day 90 mRS Ordinal |  |  | <0.001 |
| 0 (no symptoms) | 309 (25.6) | 4 (1.1) |  |
| 1 (no significant disability) | 226 (18.7) | 29 (7.9) |  |
| 2 (slight disability) | 175 (14.5) | 79 (21.5) |  |
| 3 (moderate disability) | 142 (11.8) | 44 (12) |  |
| 4 (moderate-severe disability) | 106 (8.8) | 53 (14.4) |  |
| 5 (severe disability) | 103 (8.5) | 66 (18) |  |
| 6 (dead) | 145 (12) | 92 (25.1) |  |
| Dichotomized |  |  |  |
| Day 90 Nondisabled (mRS 0-1) | 535 (44.4) | 33 (9.0) | <0.001 |
| Day 90 Independent (mRS 0-2) | 710 (58.9) | 112 (30.5) | <0.001 |

Among all the 1573 ACVD patients, day 4 mRS and day 90 mRS correlated strongly, Spearman’s rho=0.79 (*p<0.001*), in unadjusted analysis (Figure 1A). Among all 7 mRS levels, simple carry-forward of the day 4 mRS to impute a day 90 mRS matched the actual day 90 mRS with the modest observed agreement rate of 39.5%. Day 90 scores were improved (lower) in 38.8%, by 1 level in 18.0%, and by more than 1 level in 20.8%. Day 90 scores were worse (higher) in 21.6%, by 1 level in 17.0% and by more than 1 level in 4.6%. The weighted kappa, taking into account the ordered ordinal relationship of the mRS outcomes, was 0.59, demonstrating moderate above-chance concurrence. For dichotomized outcomes, simple carry-forward of the day 4 mRS to impute a day 90 mRS matched the actual day 90 mRS with considerable rates of agreement rates: mRS 0-1, 85.4%; mRS 0-2, 79.5%; fatal outcome, 88.3%. The kappa values for these dichotomized outcomes were: mRS 0-1, 0.67; mRS 0-2, 0.59; and death, 0.33.

**Figure 1A.**
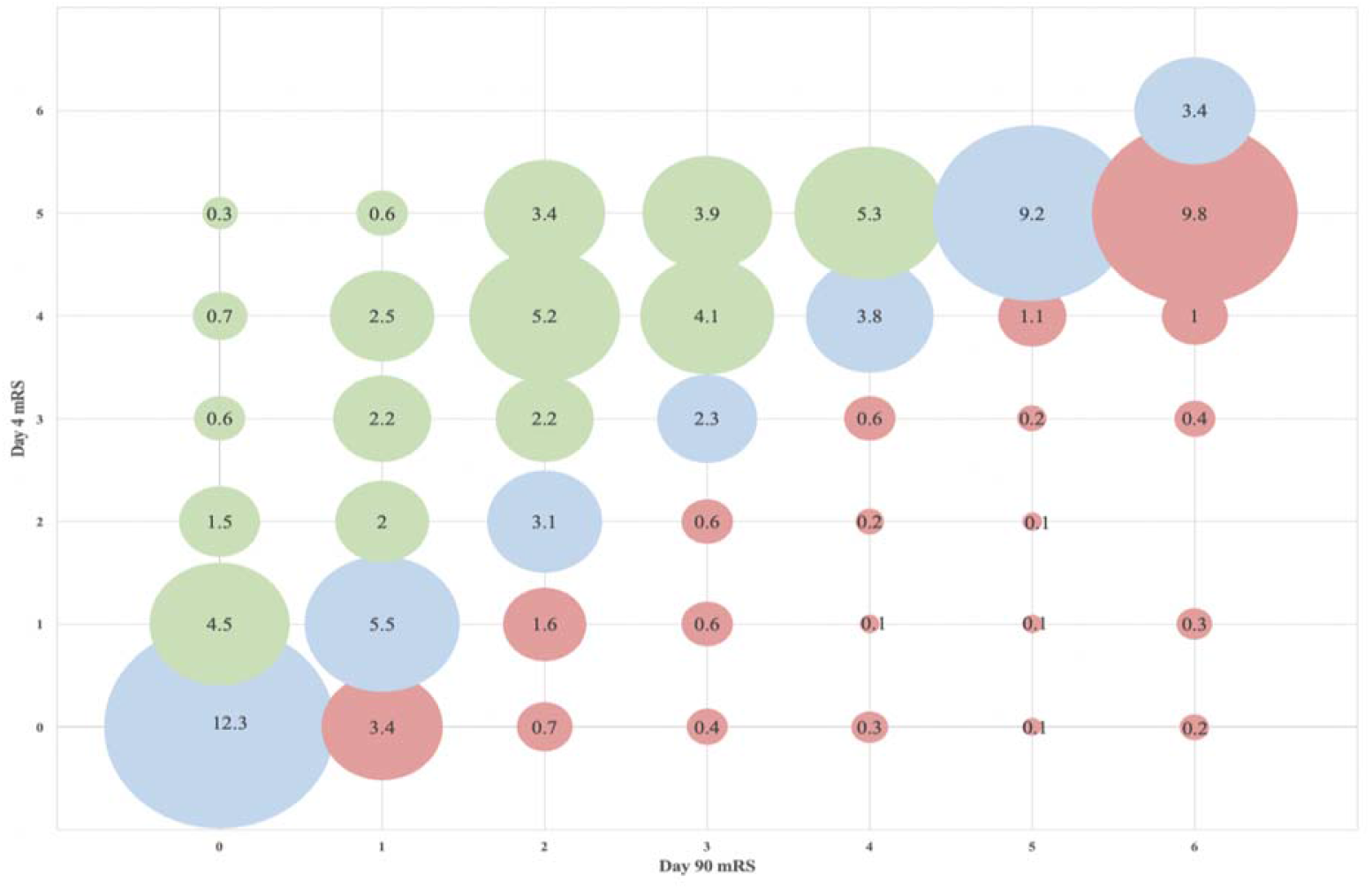
Day 4 vs Day 90 mRS, Unadjusted, Among All Acute Neurovascular Disease Patients

There were subtle differences among patients with ACI and ICH in the degree of relationship between the unadjusted day 4 and the day 90 mRS (Figure 1B, 1C). Spearman’s correlation coefficients for day 4 vs day 90 mRS were 0.76 (*p<0.001*) for patients with ACI and 0.71 (*p<0.001*) for patients with ICH. Among all 7 mRS levels, simple carry-forward of the day 4 mRS to impute a day 90 mRS matched the actual day 90 mRS in 41.3% of patients with ACI and 34% of patients with ICH. The weighted kappa was 0.71 for ACI and 0.54 for ICH. For dichotomized outcomes, simple carry-forward of the day 4 mRS to impute a day 90 mRS matched the actual day 90 mRS at the following observed agreement rates: For ACI: mRS 0-1, 85.0%; mRS 0-2, 78.0%; fatal outcome, 87.0%; for ICH: mRS 0-1, 91.8%; mRS 0-2, 75.5%; fatal outcome, 83.9%. The kappa values for these dichotomized outcomes were: For ACI: mRS 0- 1, 0.61; mRS 0-2, 0.55; and death, 0.33; For ICH: mRS 0-1, 0.31; mRS 0-2, 0.27; and death, 0.46.

**Figure 1B.**
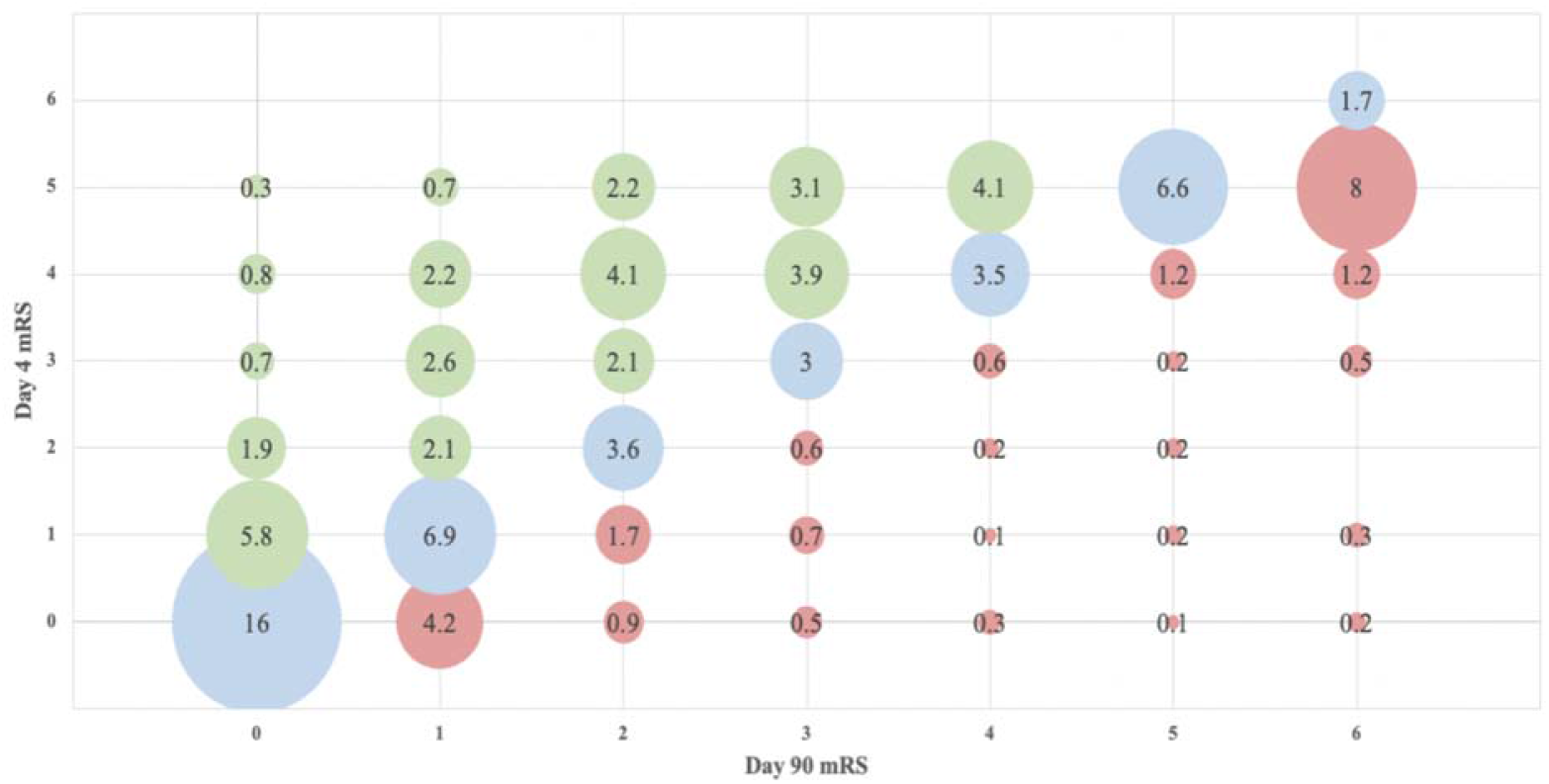
Day 4 vs Day 90 mRS in Patients with Acute Cerebral Ischemia

**Figure 1C.**
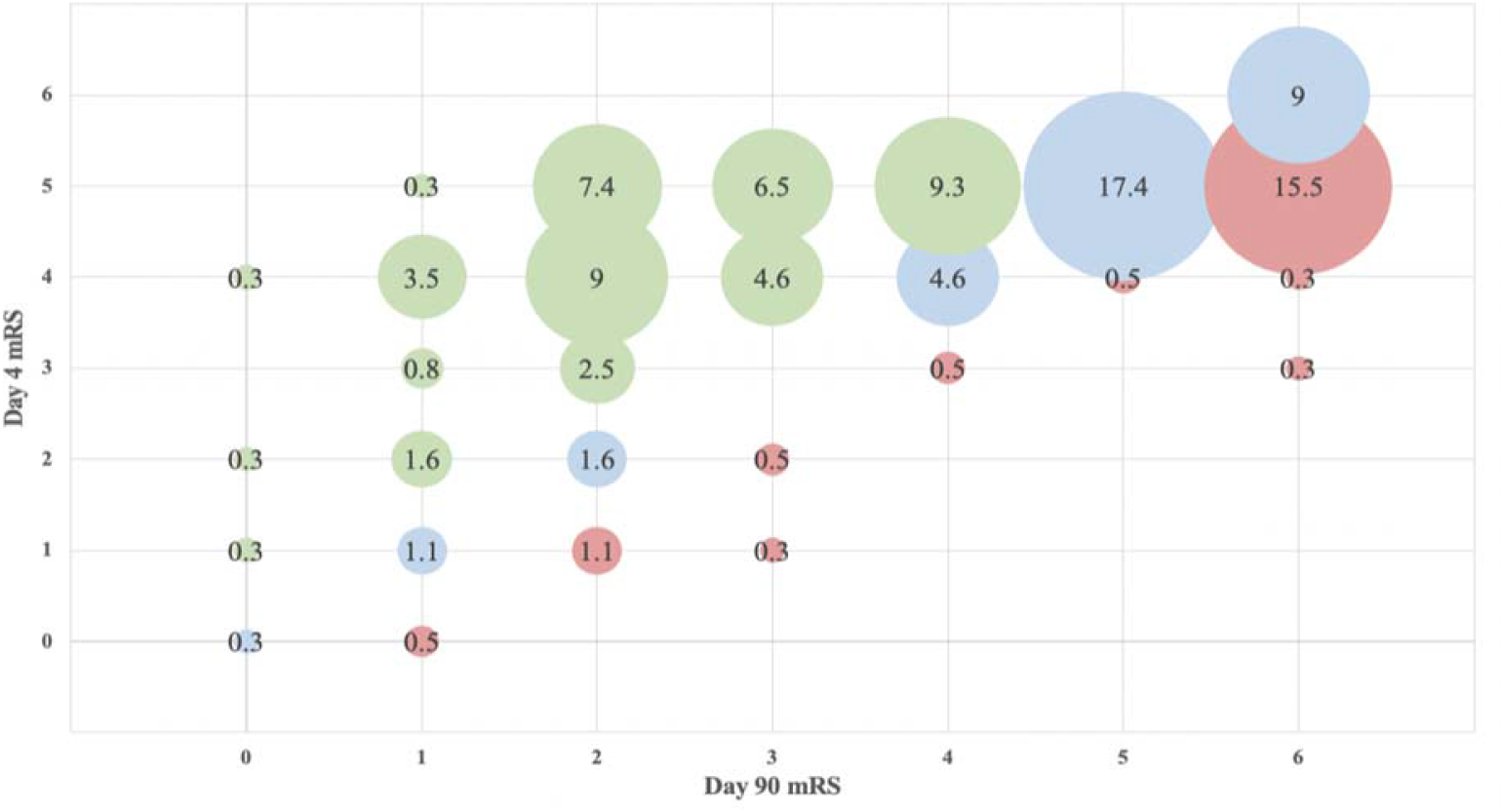
Day 4 vs Day 90 mRS in Patients with Intracranial Hemorrhage

The pattern of exact matches among the 7 levels of the mRS at day 4 and day 90 were notably different for patients with ACI and ICH. For ACI cohort, the most frequent exact match was mRS 0 at both day 4 and day 90, occurring in 16% of patients (Figure 1B), while for ICH cohort, the most frequent exact match was mRS 5 at both day 4 and day 90, occurring in 17.4% of patients (Figure 1C).

The multivariate models for predicting day 90 mRS based on day 4 mRS plus baseline covariates are shown in Table 3, for the outcomes of mRS 0-1, mRS 0-2, and ordinal mRS distribution. In addition to the clinically selected predictor variables included in all models (day 4 mRS, initial NIHSS, and age), the Akaike Information Criterion (AIC) additionally selected for incorporation into some of the models the variables of: Hispanic ethnicity, current alcohol use, hyperlipidemia, and use of tissue plasminogen activator.

Performance of the predictive models is displayed in Figure 2. Spearman’s correlation coefficients for predicted day 90 mRS vs actual day 90 mRS were 0.78 for patients with ACI and 0.80 for patients with ICH. Among all 7 mRS levels, the mRS values yielded by the multivariate imputation model incorporating day 4 mRS and baseline covariates matched the actual day 90 mRS with observed agreement rates of 78.0% for ACI and 42.3% for ICH. The weighted kappa was 0.76 for ACI and 0.78 for ICH, demonstrating substantial above-chance concurrence.

**Figure 2A.**
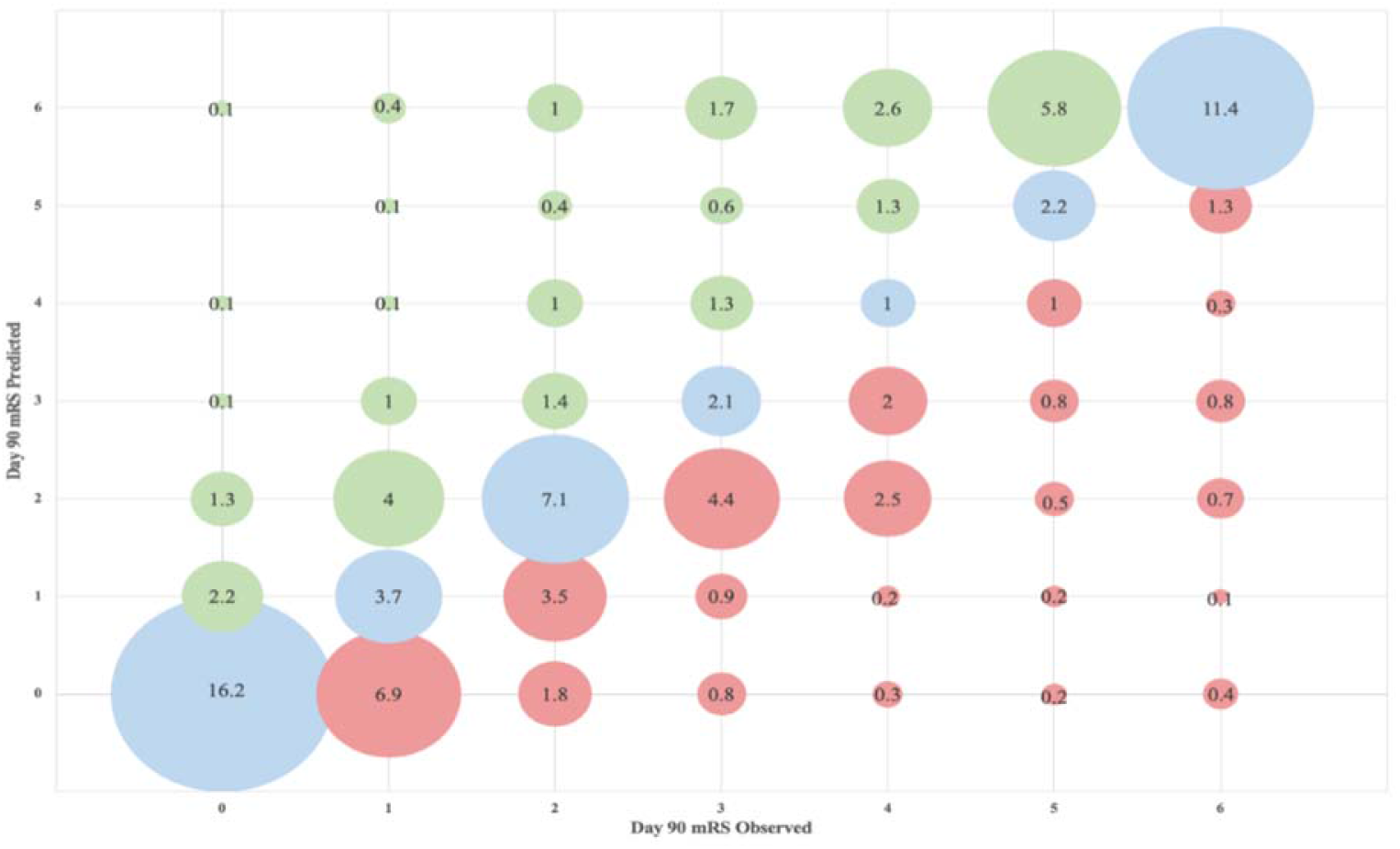
Multivariate Adjusted Day 4 vs Day 90 mRS among All Acute Neurovascular Disease Patients

**Figure 2B.**
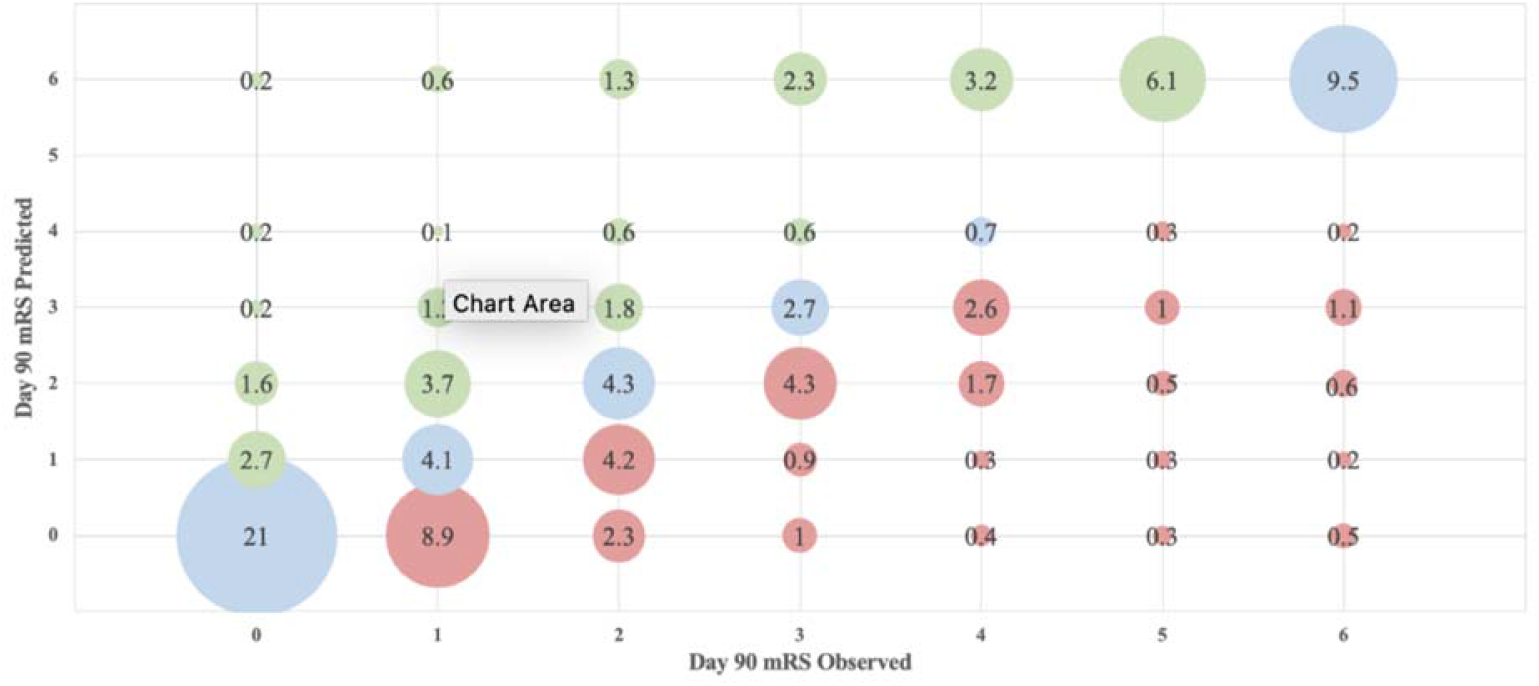
Day 4 vs Day 90 mRS in Acute Cerebral Ischemia patients

**Figure 2C.**
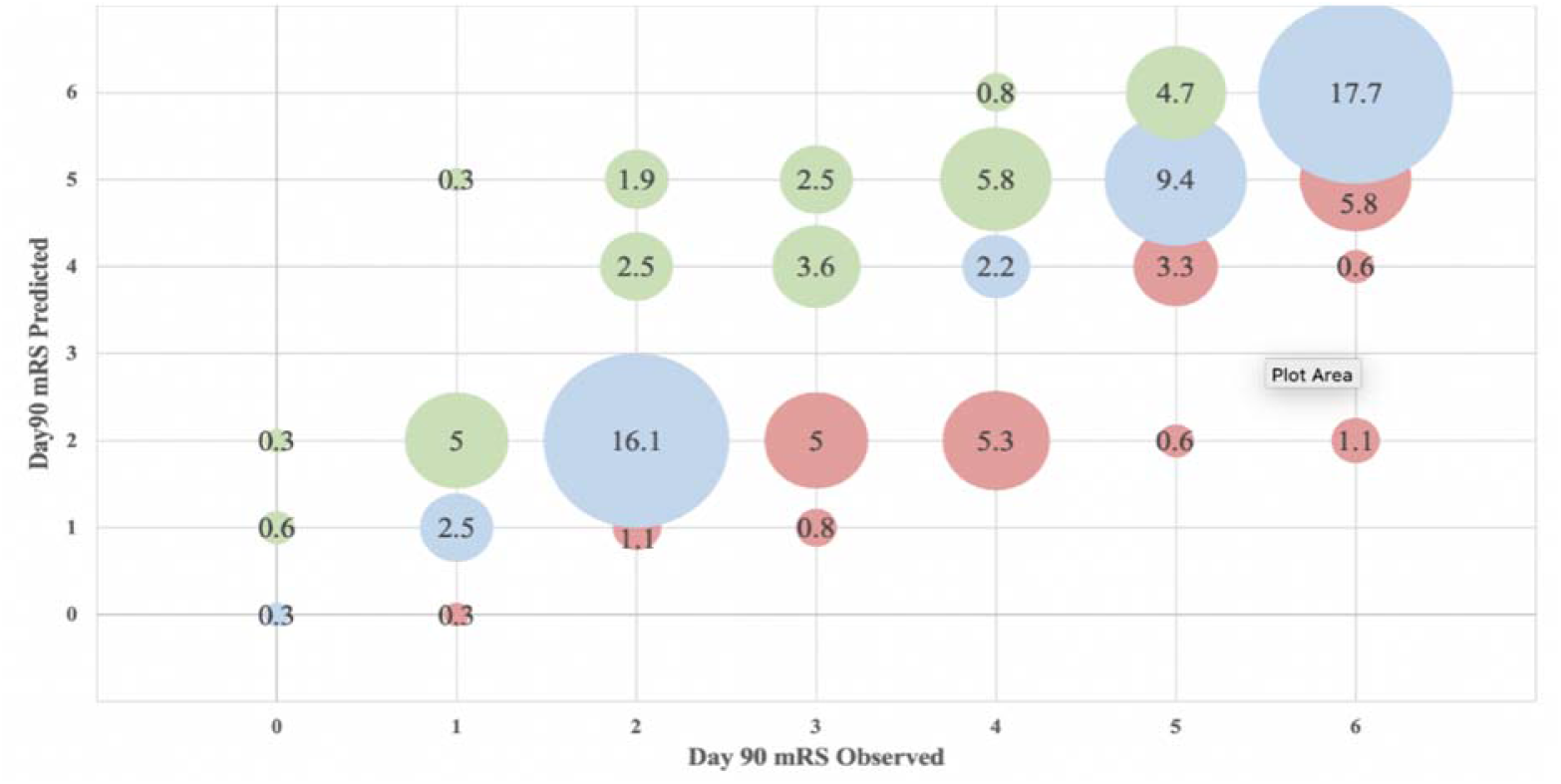
Day 4 vs Day 90 mRS in Intracranial Hemorrhage patients

For ACI, for exact matches, the predictive model incorporating day 4 mRS and other variables performed similarly to simple carry-forward of the day 4 mRS in forecasting day 90 mRS, 42.3% vs 41.3%. However, the predictive model outperformed simple carry forward in reducing directional bias in non congruent cases. With the predictive model, discrepancies showed better outcome than predicted in 25.9% of patients and worse outcome than predicted in 31.8% of patients, while simple carry-forward showed better outcome than predicted in 36.6% of patients and worse outcome than predicted in 22.1% of patients. For ICH, for exact matches, the predictive model incorporating day 4 mRS and other variables performed better than simple carry-forward of the day 4 mRS in forecasting day 90 mRS, 48.2% vs 34.0%. In addition, the predictive model outperformed simple carry forward in reducing directional bias in non congruent cases. With the predictive model, discrepancies showed better outcome than predicted in 27.9% of patients and worse outcome than predicted in 23.9% of patients, while simple carry-forward showed better outcome than predicted in 46.5% of patients and worse outcome than predicted in 19.5% of patients.

## Discussion

In this large, multicenter clinical trial, early modified Rankin Scale assessments, at day 4 post-stroke, indexed eventual 3-month mRS outcomes with good predictive accuracy. In kappa analyses, for acute cerebral ischemia, simple carry forward of the day 4 mRS to day 90 mRS showed substantial agreement for freedom-from-disability (mRS 0-1), independence (mRS 0-2), and disability level (7-level mRS), and fair agreement for mortality; multivariate predictive models incorporating day 4 mRS to predict day 90 mRS showed substantial agreement for all 4 outcomes. For intracranial hemorrhage, simple carry forward performed somewhat less well, with substantial agreement for disability level (7-level mRS) and moderate agreement for mortality, but fair agreement for freedom-from-disability (mRS 0-1) and independence (mRS 0-2); performance improved with the multivariate model, showing substantial agreement for disability (mRS 0-1), disability level (7-level mRS), and mortality and moderate agreement for independence (mRS 0-2). With simple carry forward, non congruent values between day 4 vs day 90 mRS were directionally biased, with more better-than-predicted than worse-than-predicted outcomes. But with multivariate modeling, no directional bias was present.

These findings extend prior studies. Prior investigations of the relation of early to 3- month mRS values focused solely upon patients with acute cerebral ischemia, and looked at later, day 7 and day 30, mRS assessments.^8, 9^ These studies found that mRS status at 1 week and mRS status at 1 month were strong determinants of mRS status at 3 months. In the current study, at the even earlier time point of 4 days mRS status as a sole variable predicted day 90 mRS moderately well. Comparing the three time points across studies, there is a gradation of predictive value, with mRS assessments closer to the 3-month time point showing modestly greater value in forecasting the 3-month mRS. The percent perfect agreement rates were: day 4 vs day 90 – 40%; day 7 vs day 90 – 47%; and day 30 vs day 90 – 53%. The increasing accuracy reflects patient course patterns. In the first days after acute cerebral ischemia onset, when penumbra evolution, edema, infectious complications, and other acute events are more prone to occur, patient course is variable, with fluctuations in patient status both for good or for ill. Between 1 week and 3 months, most cerebral ischemia patients do not worsen further, but some improve as neuroplasticity and brain repair restore functional capacity.^15^ Nonetheless, the current study shows that, though not quite as predictive as day 7 and day 30 mRS assessments, an mRS judgment as soon as day 4 has substantial predictive value for predicting the 3-month outcome.

For intracranial hemorrhage, to our knowledge the current study is the first to project 3- month mRS outcomes on the basis of earlier mRS observations. The day 4 mRS performed fairly well by itself in forecasting day 90 disability outcome among patients with ICH, though not as well as in patients with ACI. The patients with ICH were more severely impaired at day 4, and, from this more unstable starting point, both more often improved to a better day 90 than day outcome and more often worsened to a fatal outcome by day 90.

The predictive value of the day 4 mRS further improved when it was incorporated in multivariate models incorporating additional baseline prognostic variables, for projecting the 3- month outcomes of freedom from disability (mRS 0-1), functional independence (mRS 0-2), and mortality (mRS 6). These models determined the final outcome with high accuracy, suggesting that they are robust enough to be used for imputing missing, or not-yet-available, data in clinical trials and in quality improvement activities. Since data on patient status are always more readily available pre-discharge than post-discharge, it is advantageous to be able to project long-term outcome based on an assessment prior to patient discharge from the acute hospitalization. With the continuing reduction in the average length of stay for stroke patients in the US, the acute care in the US for ischemic stroke has now shortened substantially over the past three decades.

The one baseline variable of added value in all 3 models was the presenting neurologic deficit severity (NIHSS stroke score), consistent with the importance of neurologic impairments in determining functional disability. Baseline variables of added value in one of the models included race-ethnicity. Notably, Hispanic ethnicity was associated with a worse outcome for patients with ICH at day 90 post-stroke. Contributing factors may be less access to inpatient and outpatient rehabilitation care. Treatment with tPA was associated with worse outcomes at 3 months for patients with ACI (Table 3a). This can be explained by confounding by indication (indication bias). Compared with patients not treated with IV tPA, patients treated with tPA systematically differ in having more severe, disabling deficits at presentation.

**Table 3a.** Multivariate Predictive Models for Day 90 Freedom-from-Disability (mRS 0-1) for Patients with Acute Cerebral Ischemia and Intracranial Hemorrhage

| Acute Cerebral Ischemia |  |  | Intracranial Hemorrhage |  |  |
| --- | --- | --- | --- | --- | --- |
| Variables | Log Odds Ratio<br>(CI 0.95%) | P-value | Variables | Log Odds Ratio<br>(CI 0.95%) | P-value |
| Day 4 mRS 1 | -0.67<br>(-1.28,-0.07) | 0.03 | Day 4 mRS 1 | -18.62<br>( ) | 0.02 |
| Day 4 mRS 2 | -1.76<br>(-2.35,-2.98) | <0.001 | Day 4 mRS 2 | 0.72<br>(0.12-4.27) | 0.72 |
| Day 4 mRS 3 | -2.35<br>(-2.98,-1.73) | <0.001 | Day 4 mRS 3 | 0.15<br>(0.02-0.96) | 0.05 |
| Day 4 mRS 4 | -3.30<br>(-3.89,-2.72) | <0.001 | Day 4 mRS 4 | 0.20<br>(0.04-0.93) | 0.04 |
| Day 4 mRS 5 | -5.04<br>(-5.80,-4.27) | <0.001 | Day 4 mRS 5 | 0.02<br>(0.002-0.14) | <0.001 |
| Initial NIHSS | -0.04<br>(-1.28,-0.07) | 0.002 | Initial NIHSS | -0.14<br>(-0.25,-0.03) | 0.03 |
| Prior Stroke | 0.64<br>(-0.02,1.30) | 0.059 | Age | -0.02<br>(-0.06,0.02) | 0.28 |
| Male Gender | -0.06<br>(-0.39, 0.27) | 0.72 | Hispanic<br>Ethnicity | 1.33<br>(0.17,2.50) | 0.03 |
| TPA Treatment | 0.11<br>(-0.25,0.47) | 0.56 |  |  |  |

**Table 3b.** Multivariate Predictive Models for Day 90 Functional Independence (mRS 0-2) for Patients with Acute Cerebral Ischemia and Intracranial Hemorrhage with Functional Independence (mRS 0-2)

| Acute Cerebral Ischemia |  |  | Intracranial Hemorrhage |  |  |
| --- | --- | --- | --- | --- | --- |
| Variables | Log Odds Ratio<br>(CI 95%) | P-value | Variables | Log Odds Ratio<br>(CI 95%) | P-value |
| Day 4 mRS 1 | -0.33<br>(-1.77,1.10) | 0.64 | Day 4 mRS 0+1 | - | - |
| Day 4 mRS 2 | 0.19<br>(-1.75,2.14) | 0.84 | Day 4 mRS 2 | 0.64<br>(0.05-8.91) | 0.73 |
| Day 4 mRS 3 | -2.05<br>(-3.44,-0.67) | 0.004 | Day 4 mRS 3 | 0.52<br>(0.04-6.37) | 0.61 |
| Day 4 mRS 4 | -2.39<br>(-3.65,-1.13) | <0.001 | Day 4 mRS 4 | 0.20<br>(0.02-1.68) | 0.14 |
| Day 4 mRS 5 | -3.91<br>(-5.23,-2.59) | <0.001 | Day 4 mRS 5 | 0.03<br>(0.003-0.24) | <0.001 |
| Initial NIHSS | -0.09<br>(-0.12,-0.05) | <0.001 | Initial NIHSS | 0.91<br>(0.87-0.95) | <0.001 |
| Prior Stroke | 0.49<br>(0.51,1.51) | 0.33 | Prior Stroke | 0.09<br>(0.009-0.85) | 0.04 |
| Age | -0.05<br>(-0.07,-0.03) | <0.001 | Age | 0.93<br>(0.90-0.95) | <0.001 |
| Current Alcohol<br>Use | -0.27<br>(-0.82,0.26) | 0.31 |  |  |  |

**Table 3c.** Multivariate Predictive Models for Day 90 Level of Disability (Ordinal 7-Level mRS) for Patients with Acute Cerebral Ischemia and Intracranial Hemorrhage

| Acute Cerebral Ischemia |  |  | Intracranial Hemorrhage |  |  |
| --- | --- | --- | --- | --- | --- |
| Variables | Odds Ratio<br>(CI 95%) | P-value | Variables | Odds Ratio<br>(CI 95%) | P-value |
| Day 4 mRS 1 | 0.41<br>(0.21-0.45) | <0.001 | Day 4 mRS 1 | 0.40<br>(0.03-5.51) | 0.50 |
| Day 4 mRS 2 | 0.11<br>(0.07-0.18) | <0.001 | Day 4 mRS 2 | 0.22<br>(0.02-2.64) | 0.23 |
| Day 4 mRS 3 | 0.051<br>(0.03-0.08) | <0.001 | Day 4 mRS 3 | 0.063<br>(0.005-0.79) | 0.03 |
| Day 4 mRS 4 | 0.028<br>(0.02-0.04) | <0.001 | Day 4 mRS 4 | 0.061<br>(0.006-0.67) | 0.023 |
| Day 4 mRS 5 | 0.006<br>(0.004-0.01) | <0.001 | Day 4 mRS 5 | 0.006<br>(0.001-0.07) | <0.001 |
| Initial NIHSS | 0.95<br>(0.94-0.97) | <0.001 | Initial NIHSS | 0.92<br>(0.90-0.94) | <0.001 |
| Prior Stroke | 0.72<br>(0.50-1.03) | 0.07 | Prior Stroke | 0.26<br>(0.11-1.63) | 0.003 |
| Age | 0.98<br>(0.97-0.99) | <0.001 | Age | 0.94<br>(0.92-0.95) | <0.001 |
|  |  |  | Hyperlipidemia | 1.48<br>(0.95-2.30) | 0.10 |

Our results are important for routine clinical care and for research studies. For patients, family, and caregivers interested in long-term prognosis shortly after stroke onset, the day 4 mRS, combined with a few additional readily available variables, can provide valuable insight into likelihood of future disability. For phase 2, dose-optimization phase clinical trials using adaptive designs that require rapid feedback of treatment outcomes to drive selection of next tested dose, our findings indicate that day 4 mRS can serve as an informative early endpoint.^16^ Further, even in large trials that use day 90 assessment as the primary endpoint, our data provide an important strategy for handling the infrequent occurrence of patients being lost to follow-up after discharge from the acute care setting. The correlation of day 4 and day 90 values provides some support to the technique of last observation carried forward to fill in missing mRS outcome data, and the multivariate predictive formula strongly supports the use of multiple imputation techniques incorporating the day 4 mRS as an even more precise method for projecting missing final outcome data.^17^

This study has limitations. The study was performed in a broad and diverse population at 60 hospitals, but in a single geographic region. Replication in other geographic settings is desirable, as well as validation of the multivariate predictive models in independent populations, to confirm generalizability. Also, as studied patients were enrolled in a clinical trial, they may not be fully representative of an all-comer stroke population. The FAST-MAG trial had broad entry criteria, mitigating this concern; however, the trial did exclude patients with severe pre-stroke disability, and these patients will clearly have a different final disability outcome than those here investigated.

In conclusion, in this acute cerebrovascular disease patient cohort, assessment of global disability performed on day 4 was found to be highly informative regarding long-term, 3-month mRS disability outcome, alone, and even more strongly in combination with baseline prognostic variables. As a result, the day 4 mRS is a useful measure for imputing the final patient disability outcome in clinical trials and quality improvement programs.

## Data Availability

The datasets generated and/or analyzed during the current study are available in the ClinicaTrials.gov repository, https://clinicaltrials.gov/ct2/show/NCT00059332.

## Acknowledgments

**None**

## Figure Legend

Figure 1, 2. Crosstab tables showing proportion of patients with each of all possible pairs of day 4 and day 90 mRS scores. Blue shading indicates exact match, red shading indicates worse day 90 than day 4 mRS, green shading indicates better day 90 than day 4 mRS. A) All acute cerebrovascular disease patients, B) Acute cerebral ischemia patients, C) Intracranial hemorrhage patients.

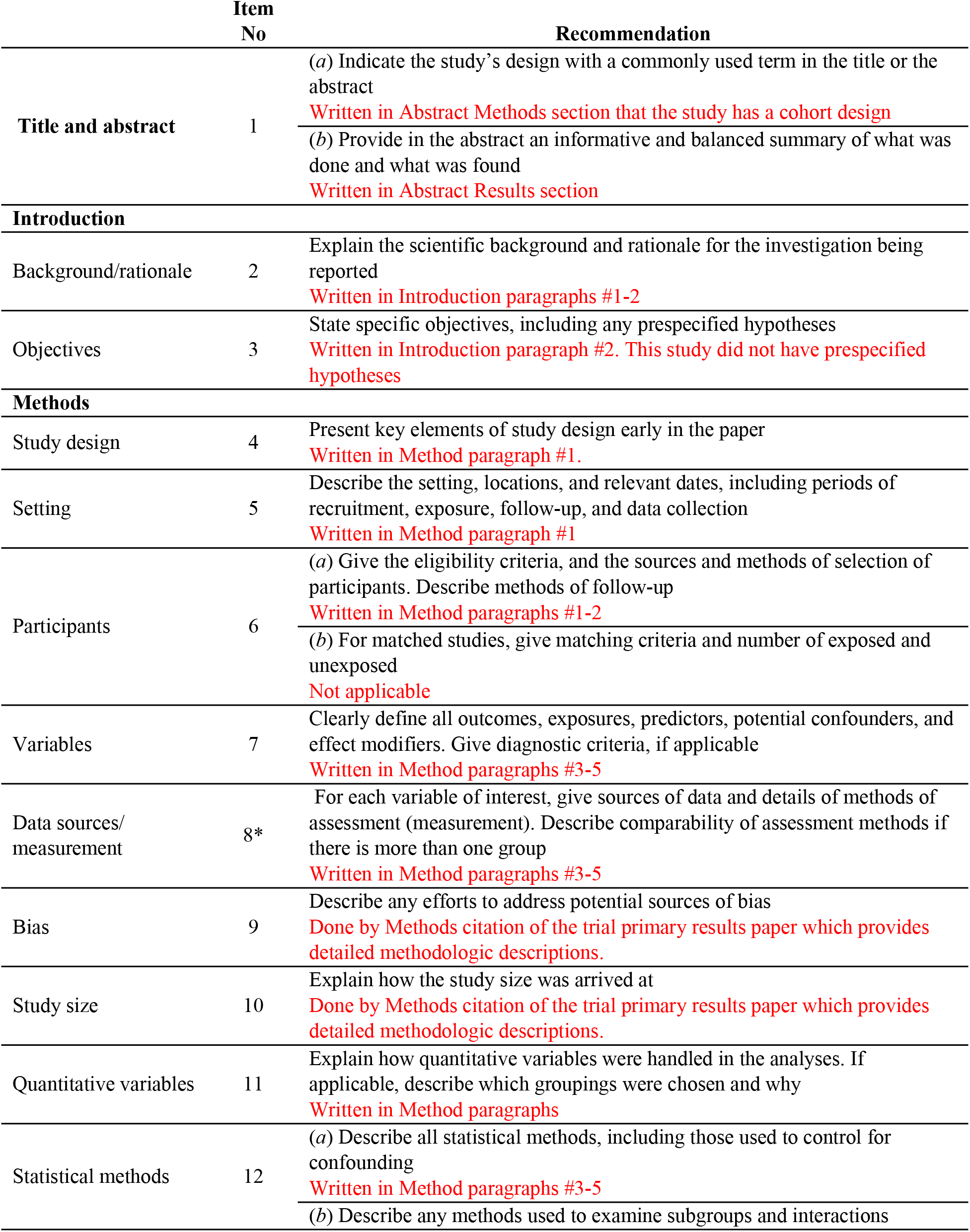
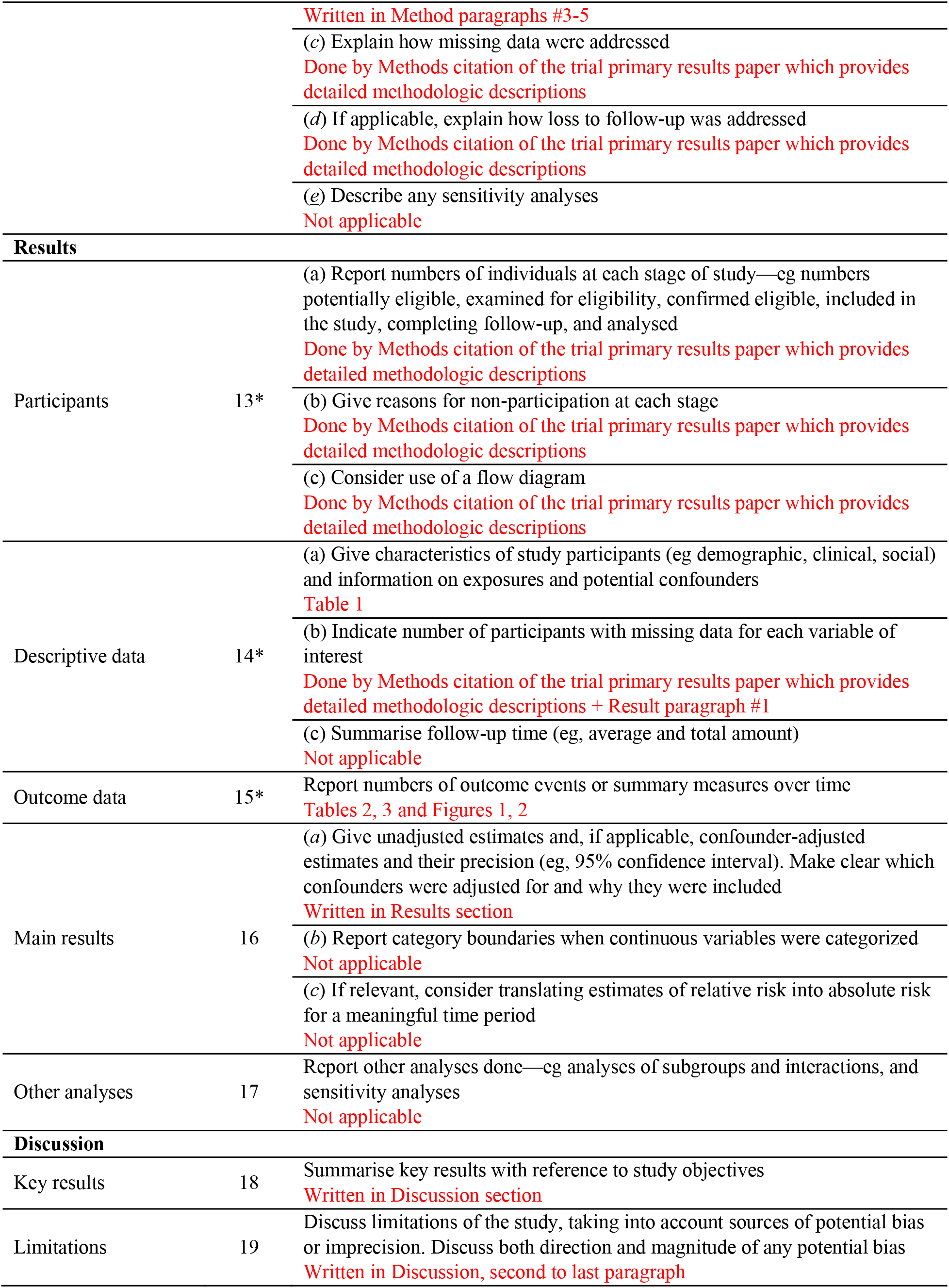
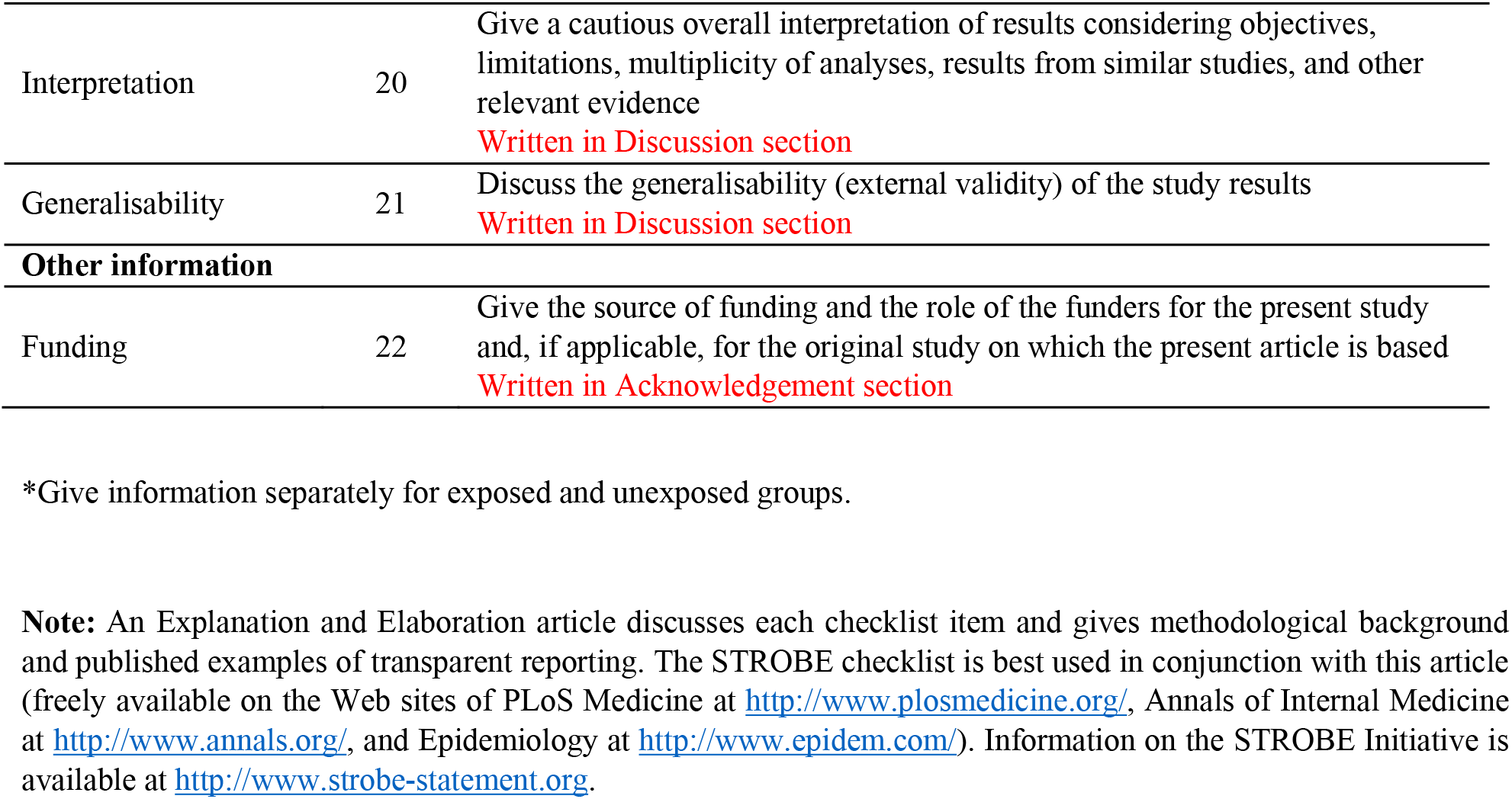
STROBE Statement—Checklist of items that should be included in reports of ***cohort studies***

